# Clinical Sensitivity and Interpretation of PCR and Serological COVID-19 Diagnostics for Patients Presenting to the Hospital

**DOI:** 10.1101/2020.06.19.20135723

**Authors:** Tyler E. Miller, Wilfredo F. Garcia Beltran, Adam Z. Bard, Tasos Gogakos, Melis N. Anahtar, Michael Gerino Astudillo, Diane Yang, Julia Thierauf, Adam S. Fisch, Grace K. Mahowald, Megan J. Fitzpatrick, Valentina Nardi, Jared Feldman, Blake M. Hauser, Timothy M. Caradonna, Hetal D. Marble, Lauren L. Ritterhouse, Sara E. Turbett, Julie Batten, Nicholas Zeke Georgantas, Galit Alter, Aaron G. Schmidt, Jason B. Harris, Jeffrey A. Gelfand, Mark C. Poznansky, Bradley E. Bernstein, David N. Louis, Anand Dighe, Richelle C. Charles, Edward T. Ryan, John A. Branda, Virginia M. Pierce, Mandakolathur R. Murali, A. John Iafrate, Eric S. Rosenberg, Jochen Lennerz

**Author notes:** These authors contributed equally (Tyler E. Miller, Wilfredo F. Garcia Beltran, Adam Z. Bard, Tasos Gogakos).

## Abstract

**Introduction:** The diagnosis of COVID-19 requires integration of clinical and laboratory data. SARS-CoV-2 diagnostic assays play a central role in diagnosis and have fixed technical performance metrics. Interpretation becomes challenging because the clinical sensitivity changes as the virus clears and the immune response emerges. Our goal was to examine the clinical sensitivity of two most common SARS-CoV-2 diagnostic test modalities, polymerase chain reaction (PCR) and serology, over the disease course to provide insight into their clinical interpretation in patients presenting to the hospital.

**Methods:** A single-center, retrospective study. To derive clinical sensitivity of PCR, we identified 209 PCR-positive SARS-CoV-2 patients with multiple PCR test results (624 total PCR tests) and calculated daily sensitivity from date of symptom onset or first positive test. To calculate daily clinical sensitivity by serology, we utilized 157 PCR-positive patients with a total of 197 specimens tested by enzyme-linked immunosorbent assay for IgM, IgG, and IgA anti-SARS-CoV-2 antibodies.

**Results:** Clinical sensitivity of PCR decreased with days post symptom onset with >90% clinical sensitivity during the first 5 days after symptom onset, 70-71% from days 9-11, and 30% at day 21. In contrast, serological sensitivity increased with days post symptom onset with >50% of patients seropositive by at least one antibody isotype after day 7, >80% after day 12, and 100% by day 21.

**Conclusion:** PCR and serology are complimentary modalities that require time-dependent interpretation. Superimposition of sensitivities over time indicate that serology can function as a reliable diagnostic aid indicating recent or prior infection.

## Introduction

While many measures to mitigate the multifactorial impact of COVID-19 are being implemented, one critical component of this strategy is the widespread testing and identification of individuals currently or previously infected by SARS-CoV-2. The delivery of effective care and mitigation of infection centers around the performance of SARS-CoV-2 diagnostic testing and the clinical interpretation of results. The lack of a full understanding of the natural history and immunopathogenesis of COVID-19 infection creates unique challenges in the implementation of diagnostic testing strategies. SARS-CoV-2 diagnostic assays have fixed technical performance metrics (e.g., sensitivity and specificity). Clinical sensitivity depends on more than technical performance and is also a function of pre-analytical variables and the disease state of the patient. Interpretation becomes challenging because the clinical sensitivity changes as the virus clears and the immune response emerges.

The goal of this study is to examine the clinical sensitivity and provide insights into the interpretation of the two most common SARS-CoV-2 diagnostic test modalities: PCR and serology. Laboratory-based diagnosis of active SARS-CoV-2 infection relies on the direct detection of virus-specific nucleic acids, most commonly obtained from the nasopharynx of infected patients. Indirect markers of infection include the detection of SARS-CoV-2 specific antibodies, generated as part of the human immune response to the virus. Serologic testing holds promise as a blood-based diagnostic aid, as a marker of viral exposure, and potentially as an indicator of protective immunity. Understanding the presence of these biomarker in relationship to one another and over the natural course of infection is required to effectively utilize these available diagnostic tests in clinical practice (1-3).

Here, we share our experience of SARS-CoV-2 PCR sensitivity and separately obtained IgM, IgA, and IgG sensitivity of an in-house enzyme-linked immunosorbent assay (ELISA) during the natural course of disease in a cohort of patients presenting to the hospital.

## Methods

### Setting and Design

The study was designed as a single-center, retrospective review of PCR results and serology data. PCR results were obtained between 3/3/2020 and 4/15/2020 and we superimposed serology data obtained from confirmed COVID-19 positive patients as part of ongoing clinical validation studies of an enzyme-linked immunosorbent assay (ELISA) for regulatory approval. The study was conducted with approval from the local Institutional Review Board.

### PCR

Nucleic acid testing was performed as part of clinical care using three emergency-use authorized real-time PCR assays. Our laboratory-developed real-time PCR assay uses the Centers for Disease Control and Prevention primers targeting regions of the N gene of SARS-CoV-22, the cobas SARS-CoV-2 Test performed on the cobas 6800 targets regions of the ORF1a and E genes, and the Xpert Xpress SARS-CoV-2 assay run on the GeneXpert Infinity targets regions of the N and E genes. The choice of testing platform was determined by access to reagents available at the time of clinical testing provided for patient care (see supplement).

### Serology

An in-house enzyme-linked immunosorbent assay was used to measure IgG, IgA, and IgM antibodies that target the SARS-CoV-2 receptor binding domain (RBD) within the spike protein. The assay has been validated within MGH clinical laboratories as a high-complexity molecular test (see supplement).

### Patient cohorts and statistical analysis

An initial PCR-query was performed to delineate clinical sensitivity over time, and analysis was restricted to patients with multiple PCR test results and at least one positive. These patients were considered confirmed COVID-19 positive and taken as true positives. In this subset, 83% were inpatients, 13% were patients from the ED, and 4% were outpatients. We calculated daily clinical sensitivity rates by dividing positive tests by all tests at each day post symptom onset and after first positive PCR-test. To estimate the time when PCR sensitivity reaches zero, we modeled a linear daily regression trend after first positive PCR-test (see supplement).

Serologic analysis of IgM, IgA and IgG status was performed in a subset of all SARS-CoV-2 PCR-positive patients and for each sample, we determined the days post symptom onset at the collection date and calculated daily sensitivity for each antibody isotype as well as sensitivity of any isotype. We plotted the sensitivity for both test modalities as percentages per overlapping 5-day leading intervals against the days since symptom onset (see supplement).

## Results

### PCR sensitivity for SARS-CoV-2 nucleic acid decreased with days post symptom onset

SARS-CoV-2 viral RNA levels decline over the course of infection(4). This decline of RNA levels clearly impacts the clinical sensitivity of PCR testing. It is not possible to determine the false negative rate of the PCR test from patients with a single PCR result. Therefore, we identified patients with multiple PCR tests who had at least one positive test result (considered true positives). The resulting dataset is composed of 624 test results from 209 patients (6.6% of all PCR-positive patients, Table 1). We compared this subset to all tested patients (Supplemental Figure 1, Supplemental Table 1) and contingency analysis of the multi-PCR vs. all single PCR-positive patient subset showed no significant differences in age, gender, and test type (Supplemental Table 1, Supplemental Figure 2). Thus, we consider the multi-PCR subset is demographically representative of our tested patient population.

**Table 1.** Demographics of cohorts used in analysis.

|  | PCR Total<br>n (%) | PCR-positive<br>n (%) | PCR Multiple Tests<br>n (%) | Serology<br>n (%) |
| --- | --- | --- | --- | --- |
| **Patients** | 11,698 | 3,163 (27) | 209 | 157 |
| **Age** |  |  |  |  |
| Median | 46 | 47 | 46<sup>a</sup> | 57<sup>a</sup> |
| Average | 47.0 | 48.0 | 48.6 | 57 |
| Range | 0 to 102 | 0 to 102 | 21 to 93 | 22 to 98 |
| **Gender** |  |  |  |  |
| Female | 6,411 (55) | 1,584 (25) | 110 (53)<sup>b</sup> | 55 (35)<sup>b</sup> |
| Male | 5,270 (45) | 1,576 (30) | 99 (47) | 102 (65) |
| Other | 22 (0.2) | 3 (14) | 0 | 0 |

To derive clinical sensitivity over disease course, we needed to define an anchor point. We chose two different anchor points: symptom onset (subjective) and first positive test (objective). First, we examined the time-course by anchoring test results to the day of symptom onset (Figure 1, Supplemental Figure 3). Clinical sensitivity remains above 90% for the first 5 days after symptom onset. In our cohort of patients presenting to the hospital, patients were admitted to the hospital a median of 8 days after symptom onset (interquartile range: 4-16 days; mean: 10 days), and between days 6-8, the clinical sensitivity of PCR ranged from 84-76%. On subsequent days the sensitivity decreases, and at day 18 the sensitivity decreases below 50%. We also compared sensitivity data from an earlier study of mildly symptomatic patients(4) and noted a steeper PCR sensitivity decline, consistent with viral levels dropping more quickly in this population (Supplemental Figure 4). This data provides sensitivity estimates at the time of presentation (e.g., a patient presents at day 10 after symptom onset). Second, we also modeled how PCR sensitivity decreases over time after the first positive PCR-test (Supplemental Figure 3 and Supplemental Figure 5). Regression modeling and extension of PCR-positivity decay (foot-point analysis) revealed that in our cohort, NP-swab specimens could stay PCR-positive beyond 20 and up to 40 days (Supplemental Figure 5).

**Figure 1:**
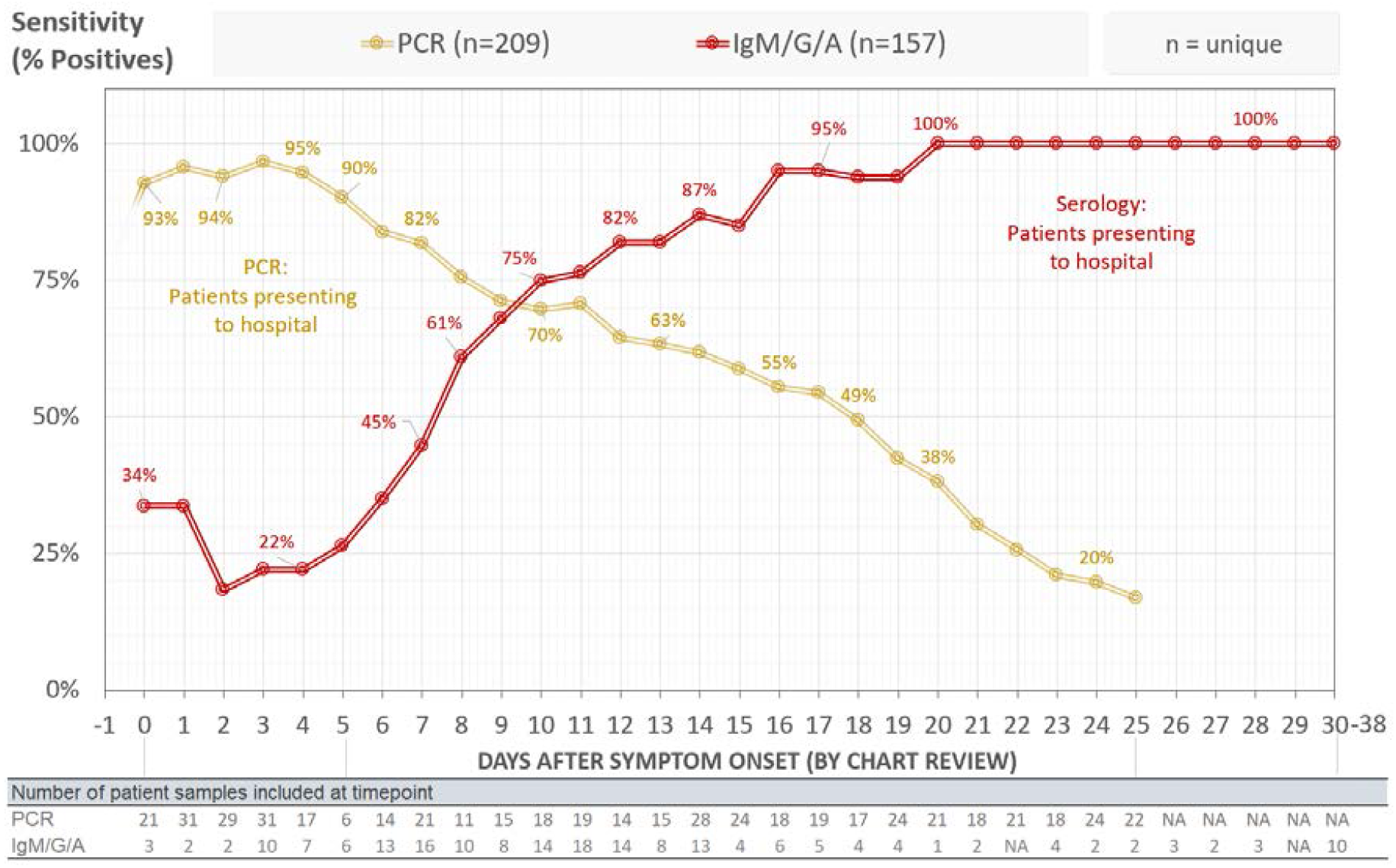
Sensitivity by assay modality over time. Blood-based serologic sensitivity in 157 patients superimposed onto NP swab PCR data from 209 patients. Results for all patient samples from initial symptom onset are plotted: PCR – 516, Serology – 588 (196 samples x 3 isotypes). Serology sensitivity is based on detection of IgM, IgG, or IgA. Sample results prior to day 0 were excluded. PCR and serology samples were obtained in largely different patient populations; therefore, sensitivities are not additive. Data is plotted as 5-day moving average against the days since symptom onset. NA: none assessed.

### Serological assay sensitivity increases with days post symptom onset

Seroconversion is also a dynamic response to the virus and assay sensitivity changes over time. To assess the sensitivity of our serology assay over time, we tested for IgM, IgG, and IgA antibodies against the receptor binding domain (RBD) of SARS-CoV-2 spike protein in a 157 SARS-CoV-2 PCR-positive patients using an in-house ELISA (Table 1). For some patients, we were able to assess serology at multiple time points (n=591 total isotype tests on 197 total specimens). Anchoring our serologic results to days after symptom onset shows that seroconversion starts as early as symptom onset, is detectable in 50% of subjects after day 7, and continues to increase with >80% of patients showing seropositivity after day 12 (Table 2, Figure 1, Supplemental Figure 4, Supplemental Table 2). In the subset of patients with multiple serological tests we saw isotype switching occur as short as 1-4 days, consistent with recent reports(5). Of note, we detected IgA prior to IgM or IgG in a number of individuals, with 2 subjects having only detectable IgA within 3 days of symptom onset. We also documented cases of IgG positivity prior to IgM or IgA, highlighting the utility of measuring seroconversion using all 3 isotypes (Supplemental Table 3, Supplemental Figure 6). The superimposition of serologic sensitivities with PCR sensitivities shows that, after day 7, seroconversion is a reliable diagnostic aid indicating recent or prior infection (Figure 1).

**Table 2.**
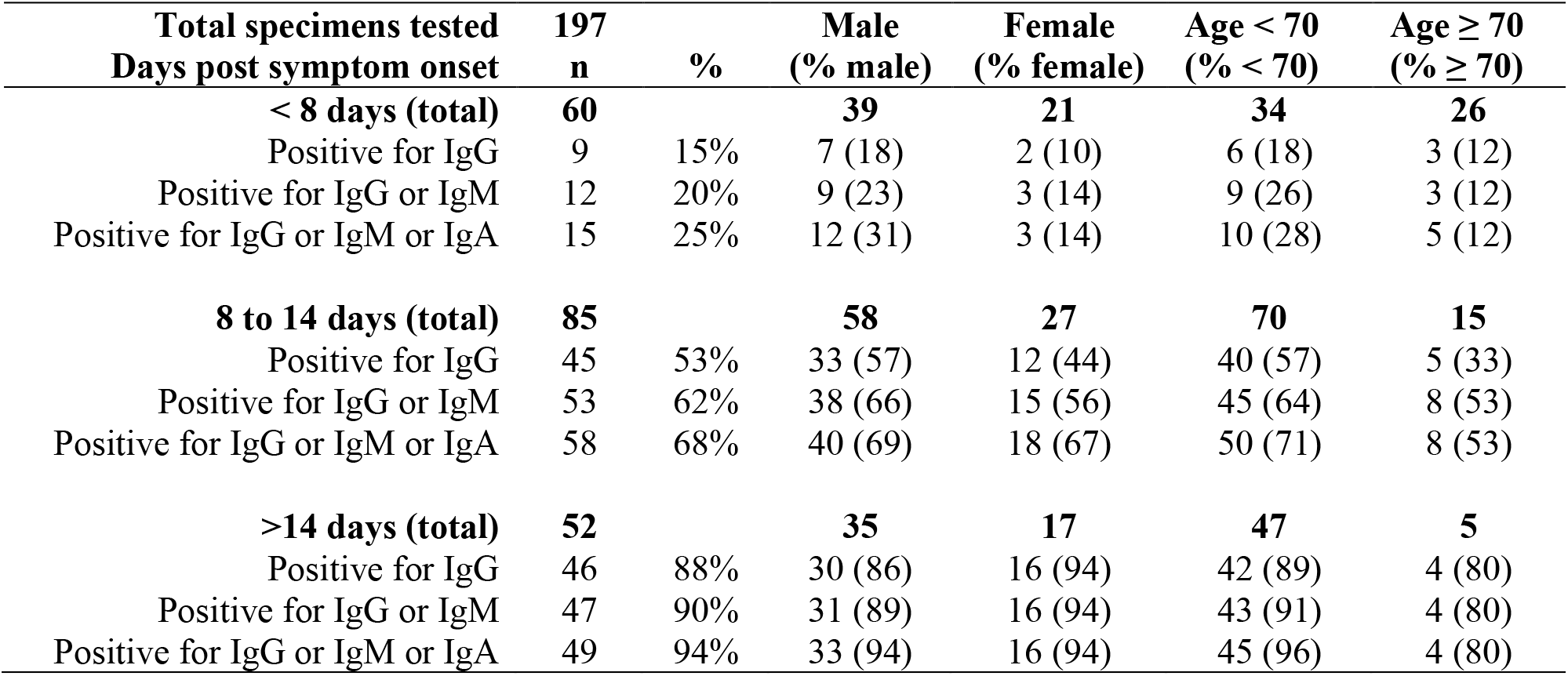
Sensitivity of anti-SARS-CoV-2 serology by isotype, age, gender, and days post symptom onset.

## Discussion

We present the dynamic clinical sensitivity of SARS-CoV-2 PCR and our serology platform in patients presenting to the hospital. We performed this single-center, retrospective analysis to share early real-world evidence that can inform interpretation of PCR and serologic testing. We focused on assessing the clinical utility of both modalities in conjunction by direct superimposition of both sensitivity time courses. The direct superimposition shows that serology can function as a reliable diagnostic aid indicating recent or prior infection – in particular at times when PCR sensitivity is lower than 70%. Our findings emphasize that understanding the specific sensitivity kinetics of both modalities is paramount for interpretation and effective utilization of SARS-CoV-2 diagnostics.

There is considerable interest in moving SARS-CoV-2 diagnostics to evidence-based principles (6-9). While clinicians await formal guidance from large, prospective, multi- center studies – which will be challenging during the ongoing pandemic – there is considerable uncertainty surrounding SARS-CoV-2 diagnostics in clinical practice (3, 10-13). Using available published data (3-7, 12, 14) and data presented here from our hospital, we offer the following five diagnostic principles for consideration:

1. **In symptomatic patients, all interpretations are anchored on days post symptom onset**. Understanding performance characteristics of SARS-CoV-2 diagnostics over the course of the infection is key to interpretation of results. We provide two distinct approaches to anchor interpretation over the course of infection: subjective (date of symptom onset) and objective (first PCR positive result). Both approaches are valid and have limitations. For example, the quality of patient histories is variable and in many cases the day of symptom onset is unknown or cannot easily be reconstructed. As a practical suggestion and whenever available, we place the date of symptom onset in the front page of the (electronic) medical record.
2. **PCR is the diagnostic gold standard during acute infection**. PCR-testing using consensus primers has an estimated specificity of >99% (15). Based on early reports from Wuhan(16) the overall clinical sensitivity is reported around 70%. We found a clinical sensitivity around 95% in the first five days after symptom onset and although PCR is an imperfect standard concurrent IgM/IgA/IgG antibody assessment in the first 5 days post-symptom onset does not significantly aid in rendering a current diagnosis, and at no point during active infection should serology replace PCR for diagnosis.
3. **Clinical sensitivity of PCR decreases with days post symptom onset**. In clinical practice many symptomatic patients present to medical care after day 1 of symptom onset. Our data show PCR sensitivity decreases with days post-symptom onset (Figure 1) or days post first PCR-positive test result. Notably, some patients may have an initial PCR-negative result at presentation (Supplemental Figure 3). Our data also indicate that severely ill patients (many patients in our cohort) remain PCR-positive for a longer period than mildly ill patients (patients in the Wölfel et al.(4) cohort). Both time and disease severity may be key elements for the interpretation of PCR results.
4. **Serological assay sensitivity increases with days post symptom onset**. In our cohort and with our assay, clinical sensitivity of serologic testing surpasses that of PCR after days 8-10 post symptom onset. Remarkably, we find seroconversion does not follow the typical kinetics of IgM antibodies followed by class-switched IgG and IgA antibodies. Rather, all appear simultaneously at a cohort level, with IgG or IgA seropositivity preceding IgM responses in some cases. Supporting this are other studies that report overall low IgM responses to SARS-CoV-2 that are often preceded by IgG(14, 17). These data highlight the benefit of measuring all three anti-SARS-CoV-2 antibody isotypes to maximize sensitivity. In the case of SARS-CoV-2, it is unknown for how long IgM, IgA or IgG antibodies remain detectable after infection. It is important to note that a positive serologic result for IgM, IgA and/or IgG does not conclusively indicate that a patient’s presenting symptoms are due to a current SARS-CoV-2 infection.
5. **Negative results do not completely preclude SARS-CoV-2 infection**. Ruling out SARS-CoV-2 infection remains challenging. Supplementing PCR results with serologic assessment can increase sensitivity (serology as a diagnostic aid). However, our data clearly shows that there is a window period (day 6-12 from symptom onset) when clinical sensitivity of PCR and serological assays are below 90%. In a symptomatic patient, if multiple PCR tests are negative and serological results after 8-12 days are also negative, we believe the likelihood of active SARS-CoV-2 infection to be low. Clinical judgement is needed in this situation as there are rare scenarios where PCR negativity may be due to disease at a different anatomic site and/or serologic negativity may be due to an immunocompromised state.

Limitations in our study include relatively small numbers, a retrospective design, and selection bias due to the specific setting and testing practice. We evaluated symptomatic, mostly hospitalized patients and we cannot derive recommendations for asymptomatic or mildly symptomatic patients from our data. Due to limited availability of tests and time constraints during an ongoing pandemic, we do not have daily samples. Date of symptom onset is not consistently available, subjective, and affected by recall biases – yet, it represents a useful anchor point for disease time course in symptomatic patients. Notably, the eclipse period ranges from 2-14 days (18-21) and some patients already mounted a serologic response at the time of presentation, which can be taken as an argument for the early y-axis deviation from zero in the serology curve and a confirmation of date of symptom onset as an imperfect marker. We caution that the presented serology data are specific to our ELISA, and we cannot extrapolate to anti-SARS-CoV-2 antibody responses in general. Nonetheless, other publications indicate that the time courses are comparable (14, 17, 22). Our serological studies measured antibodies to the RBD of SARS-CoV-2. We chose this viral antigen because of its specificity to SARS-CoV-2, and because anti-RBD antibodies are typically neutralizing. Plaque reduction neutralization tests are the gold-standard for assessing neutralizing ability (23-26), and ongoing studies are in progress to confirm anti-RBD antibodies are neutralizing in SARS-CoV-2 infection (27). Some commercially available assays measure the more abundant nucleocapsid protein, which may increase sensitivity to detect a serologic response early in the course of infection, therefore shifting the seroconversion curve to the left. However, antibodies to nucleocapsid protein are unlikely to provide protective immunity as nucleocapsid protein is inaccessible to antibodies in an intact virus. Therefore, serology results also likely depend on the specific antigen being employed in testing. Finally, our multi-PCR cohort and serologic patient cohort are largely non-overlapping (n=20/209) PCR-positive patient. To enable additive sensitivity calculations from combined PCR and serology assays, prospective and systematically obtained repeated parallel PCR and quantitative serologic data will be necessary.

Determining the clinical sensitivity of SARS-CoV-2 diagnostic tests is an area that does not have time to wait for full, exact analysis during an ongoing pandemic. Our real-world data outline the strengths and weaknesses of two SARS-CoV-2 test modalities over the natural course of infection. We hope these data, in conjunction with the 5 diagnostic principles for consideration, can contribute to effective utilization and interpretation of COVID-19 related laboratory data for patient care.

## Data Availability

Deidentified, raw data can be provided by request.

## Acknowledgments

The authors thank all patients and laboratory staff for making this study possible. CR3022 reference sequences, CR3022-IgA and CR3022-IgM isotypes, and ELISA protocols were kindly provided by Stephanie Fischinger, Caroline Atyeo, and Matthew Slein from the laboratory of Dr. Galit Alter (Ragon Institute). We thanks Yael Heher MD for excellent discussions. ELISA protocols were developed and optimized with the laboratory of Dr. Alejandro Balazs (Ragon Institute). This work is also in part supported by NIH (RO1 CA225655) to J.K.L, Centers for Disease Control and Prevention U01CK000490 (E.T.R., R.C.C.), NIGMS training grant T32 GM007753 (B.M.H., T.M.C), NIH R01 AI146779 (A.G.S.) The content is solely the responsibility of the authors and does not necessarily represent the official views of the National Institute of Health or any other organization.

